# Postingestive reinforcement is conserved in obesity and after bariatric surgery

**DOI:** 10.1101/2023.07.17.23292625

**Authors:** Gabriela Ribeiro, Ana B. Fernandes, Francisco P.M. Oliveira, João S. Duarte, Maria Oliveira, Clotilde Limbert, Rui M. Costa, Durval C. Costa, Albino J. Oliveira-Maia

**Affiliations:** Champalimaud Research & Clinical Centre, Champalimaud Foundation, Av. de Brasília, Doca de Pedrouços, 1400-038 Lisboa, Portugal; Lisbon Academic Medical Centre PhD Program, Faculdade de Medicina, Universidade de Lisboa, Avenida Professor Egas Moniz, 1649-028 Lisboa, Portugal; NOVA Medical School, Faculdade de Ciências Médicas, NMS, FCM, Universidade NOVA de Lisboa, Campo Mártires da Pátria 130, 1169-056 Lisboa, Portugal; Department of Endocrinology, Centro Hospitalar de Lisboa Ocidental, Rua da Junqueira, 126 1340-019 Lisboa, Portugal; Department of Neuroscience and Neurology, Zuckerman Mind Brain Behavior Institute, Columbia University, New York, NY, USA; Allen Institute Seattle, WA, USA

**Keywords:** Postingestive reinforcement, dopamine D2-like receptors, obesity, gastric bypass, sleeve gastrectomy, sweet taste.

## Abstract

Postingestive reward contributes to acquisition of food preferences, mediated by striatal dopamine, with assessment in humans, while challenging, suggesting blunted brain responses to postingestive nutrient stimulation in obesity. To perform postingestive conditioning in humans, we used carboxymethylcellulose, a food thickener, to optimize conditions where maltodextrin, an insipid carbohydrate, was not detectable by sensory cues (n=159). In the resulting Flavour Nutrient Conditioning protocol using flavoured yoghurts, where one flavour was paired with maltodextrin (+102 Kcal, CS^+^) and another with carboxymethylcellulose (+1.8 Kcal, CS^-^), we found that in healthy volunteers (n=52), preference for CS^+^ increased after conditioning, when assessed according to intake, with no effects on pleasantness scores. This protocol and [^123^I] IBZM SPECT, to assess availability of striatal dopamine D2-like receptors (DD2lR) were applied in a clinical study (n=61) with pre-bariatric candidates with obesity, weight-stable patients after surgery, and an additional group of healthy controls. Conditioning was conserved among participants in the clinical study, and did not differ significantly between the 3 groups. However, striatal DD2lR availability was reduced in patients from the obesity group, when compared both to healthy volunteers and the surgical group. Importantly, in exploratory analyses, DD2lR availability was strongly correlated with conditioning strength, as well as a measure of restrained eating, but only in patients with gastric bypass. These results suggest that postingestive reinforcement, while conserved in obesity and after bariatric surgery, may be associated to post-surgery recovery of central dopaminergic homeostasis and to changes in feeding behaviour after gastric bypass.

## Introduction

Postingestive signals about the energy content of food are crucial determinants of food selection, in addition to explicit sensory cues^1–3^. One extensively researched model of how rodents acquire preferences according to postingestive nutrient value is Flavour Nutrient Conditioning (FNC), with induction of a conditioned preference for an oral flavour, resulting from repeated pairings with the postingestive consequences of a nutrient^1^. FNC protocols have also been tested in humans, despite considerable methodological challenges^4^. Bland carbohydrates such as maltodextrin^5^ have been used to minimize sensory cues and thus isolate postingestive consequences. For example, FNC protocols using flavoured beverages either with added calories from maltodextrin (+112.5 Kcal) or non-caloric controls, resulted in modest increases in liking ratings for the former, but not the latter^6^, and absent effects when either higher or lower doses of maltodextrin were used^7^. However, despite its bland sensory properties, maltodextrin can be detected at different concentrations^5, 8^, raising concerns about the actual isolation of its postingestive effects. Nevertheless, in rodents, there is evidence that the postingestive consequences of sugars can induce ingestive preferences, even in the absence of orosensory input^9, 10^. Furthermore, we have shown that the postingestive effects of sucrose sustain food-seeking behaviours, that are dependent on the activity of dopaminergic neurons in the ventral tegmental area (VTA), and at least partially mediated by the hepatic branch of the vagus nerve^11^.

Obesity has been associated with altered reward-related feeding behaviour^1, 12, 13^ and brain changes that may be associated to overeating, such as lower striatal dopamine D2-like receptors (DD2lR) availability^14^ (for review, see Ribeiro G et al.,^15^). Importantly, a recent paper suggested that brain responses to postingestive administration of nutrients, including striatal responses and dopamine release, are impaired in patients with obesity, and not recovered after moderate, diet-induced weight-loss^16^. However, direct comparisons between patients with obesity and lean participants to address the behavioural effects of postingestive nutrients, as well as the impact of bariatric surgery in the former, are lacking. Indeed, bariatric surgery, currently the most effective treatment for severe obesity^17–19^, potentially normalizes obesity-related features of reward-related feeding behaviour^20–24^, with reports of food preference shifts after surgery^20, 21, 23, 25^, from energy-dense and palatable foods (e.g., rich in fats and sugars) towards less energy-dense options (e.g., fruits and vegetables). However, the mechanisms underlying these changes are not fully understood^1^. Changes in food reward processing, including postingestive reward, are potentially involved, but remain unexplored.

Here, we hypothesized that postingestive reward, as measured in a conditioning paradigm, is impaired in obesity, when compared with a healthy and lean sample, and is recovered by bariatric surgery. To address this hypothesis, we first developed a novel FNC protocol in healthy volunteers, fully addressing potential confounders from the orosensory cues provided by maltodextrin. Postingestive conditioning strength obtained in this optimized protocol was then compared between patients with obesity either prior to or after bariatric surgery, and healthy volunteers, all of whom were also assessed with [^123^I] IBZM SPECT to explore potential associations between postingestive conditioning and DD2lR availability. Additional aims included testing the differential impact of gastric surgery type, i.e., gastric bypass and sleeve gastrectomy, on postingestive reinforcement as well as DD2lR availability.

## Results

### Study overview

This study was conducted in 272 participants, tested in one of 3 main experiments. Conditions for optimal use of maltodextrin in FNC, with identification and masking of the orosensory cues resulting from consumption of maltodextrin solutions, were tested in 159 healthy participants (‘Maltodextrin optimization’ group), allowing for definition of a controlled protocol for FNC. This was applied in 52 other healthy volunteers (‘FNC development’ group), to assess behaviour in the FNC protocol and perform further optimization. In a final group of 61 participants, to compare patients with obesity either before or after bariatric surgery and a new group of healthy controls, data was collected with the final FNC protocol and nuclear medicine imaging of DD2lR availability. Please see **Table 1** for a detailed demographic and clinical description of participants.

**Table 1.**
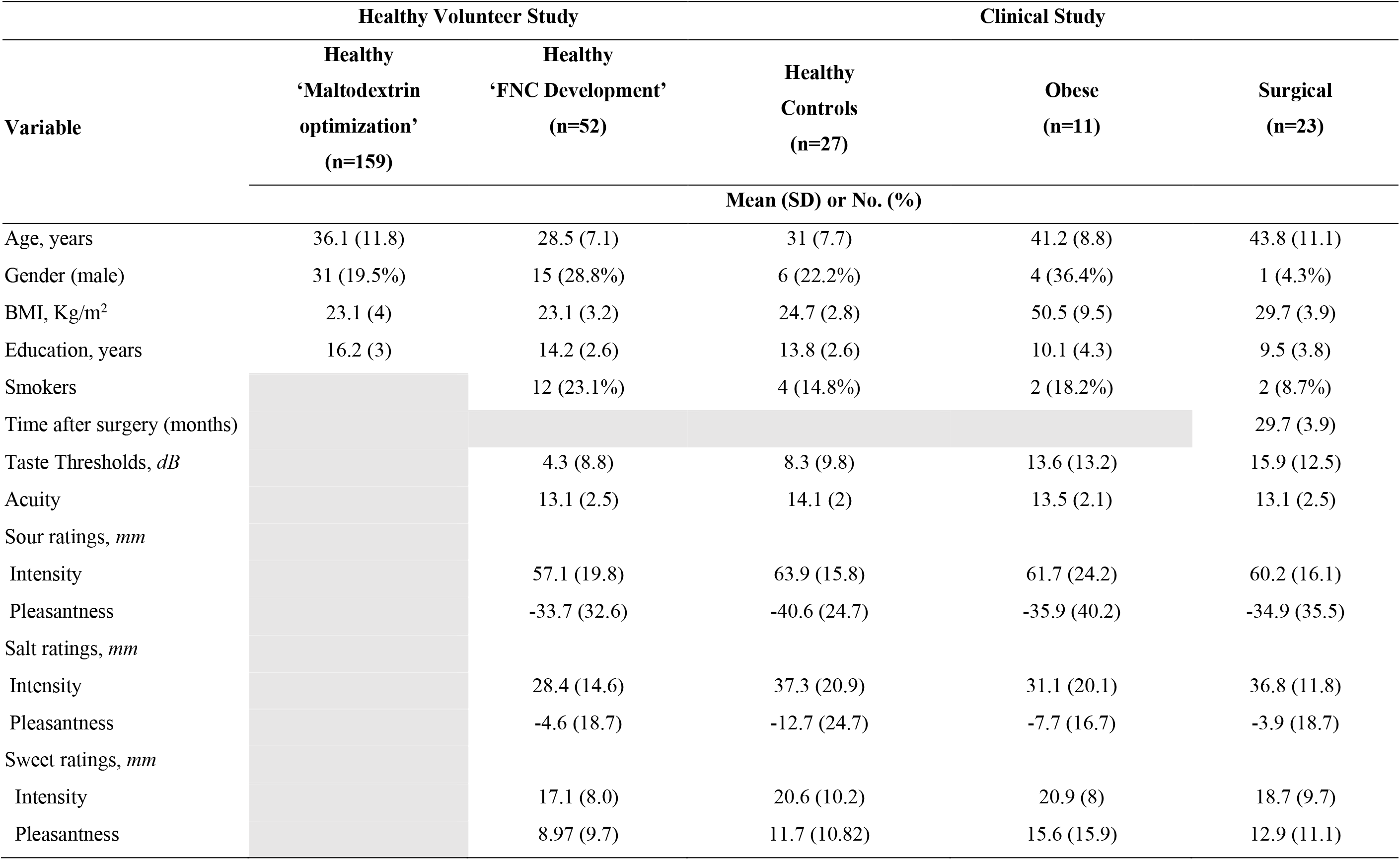
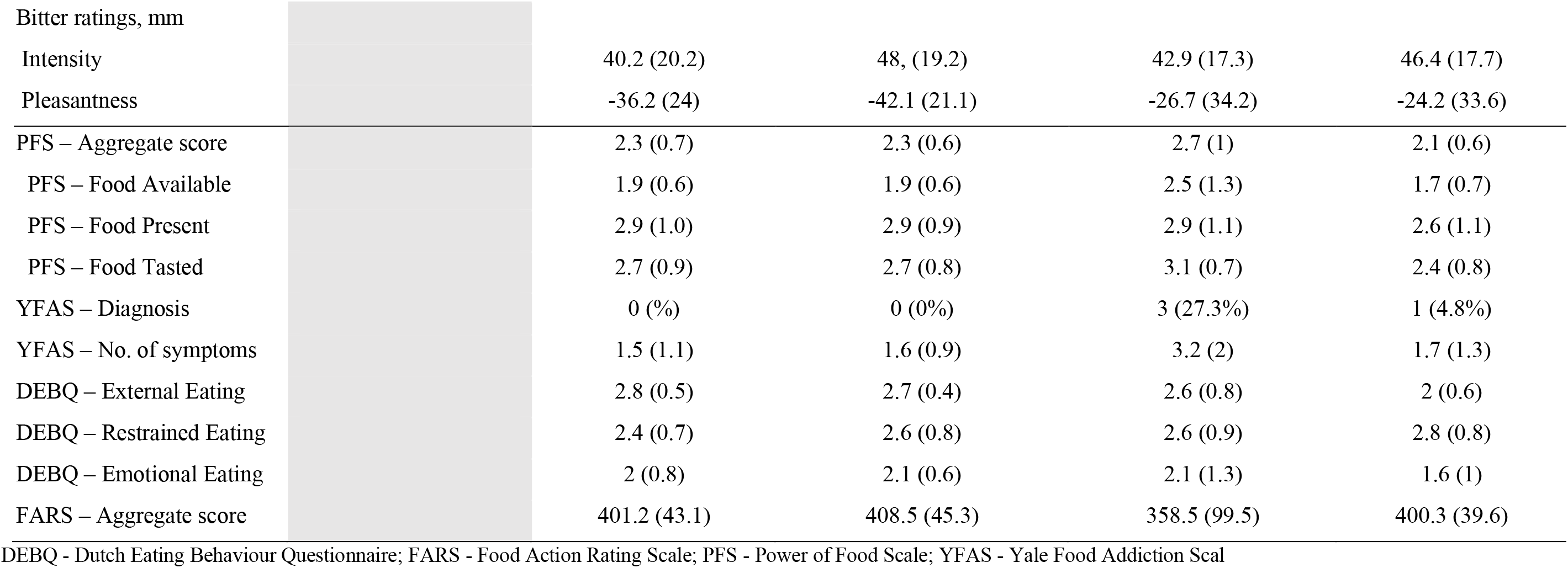
Demographic, gustatory and psychometric measures of feeding behavior in healthy subjects.

### Maltodextrin is identified through orosensory cues

While maltodextrin is typically used in FNC protocols as a source of calories due to insipid taste, we had evidence from preliminary qualitative experiments that sweetness and texture could be cues for detection of maltodextrin solutions. We thus developed a series of experiments to determine conditions in which maltodextrin, when dissolved in low-fat yoghurt sweetened with sucralose at 0.01% (w/v), would not be discriminated from the base low-fat yoghurt solution without maltodextrin added. We started by testing different maltodextrin dextrose equivalents (DE), since there are reports of increasing sweetness for higher DE^26^. Indeed, we found that yoghurt intensity ratings, normalized relative to the base yoghurt solution without maltodextrin, varied according to maltodextrin DE (4-7, 13-17 and 16.5-20; F_(2,72)_=3.4, *P*=0.04), but not concentration (17%, 25% and 33% w/v; F_(2,36)_=0.5, *P*=0.6) nor their interaction (F_(4,72)_=0.7, *P*=0.6; n=39; **Figure 1 A)**. We also found that, at the lowest maltodextrin DE (4-7) and concentration (17%), in 3-alternative forced-choice (3-AFC) tests with one or two stimuli consisting of maltodextrin yogurt, and the remaining (respectively two or one) of base yogurt, participants very easily identified maltodextrin above chance level (*P*<0.0001; n=18; **Figure 1 B**). Since all yogurts were similarly sweetened with sucralose, and we had evidence from preliminary experiments that texture could be the major cue for maltodextrin detection, we then repeated 3-AFC tests, but added carboxymethylcellulose (CMC; 0.4% w/v), a flavourless and low energy food thickener, to the base yoghurt solution. While 25% and 33%, maltodextrin was still identified significantly above chance level (25.0%, *P*=0.02, n=22; 33.0%, *P*=0.001, n=25), the lowest concentration (17%) maltodextrin yoghurt was not discriminated from CMC yoghurt solutions (*P*=0.22, n=23). This was conserved when yoghurt were flavoured, as planned for FNC experiments (*P*=0.13, n=32; **Figure 1 C**). Thus, in subsequent experiments, we used the contrast 17% maltodextrin/0.4% CMC.

**Figure 1.**
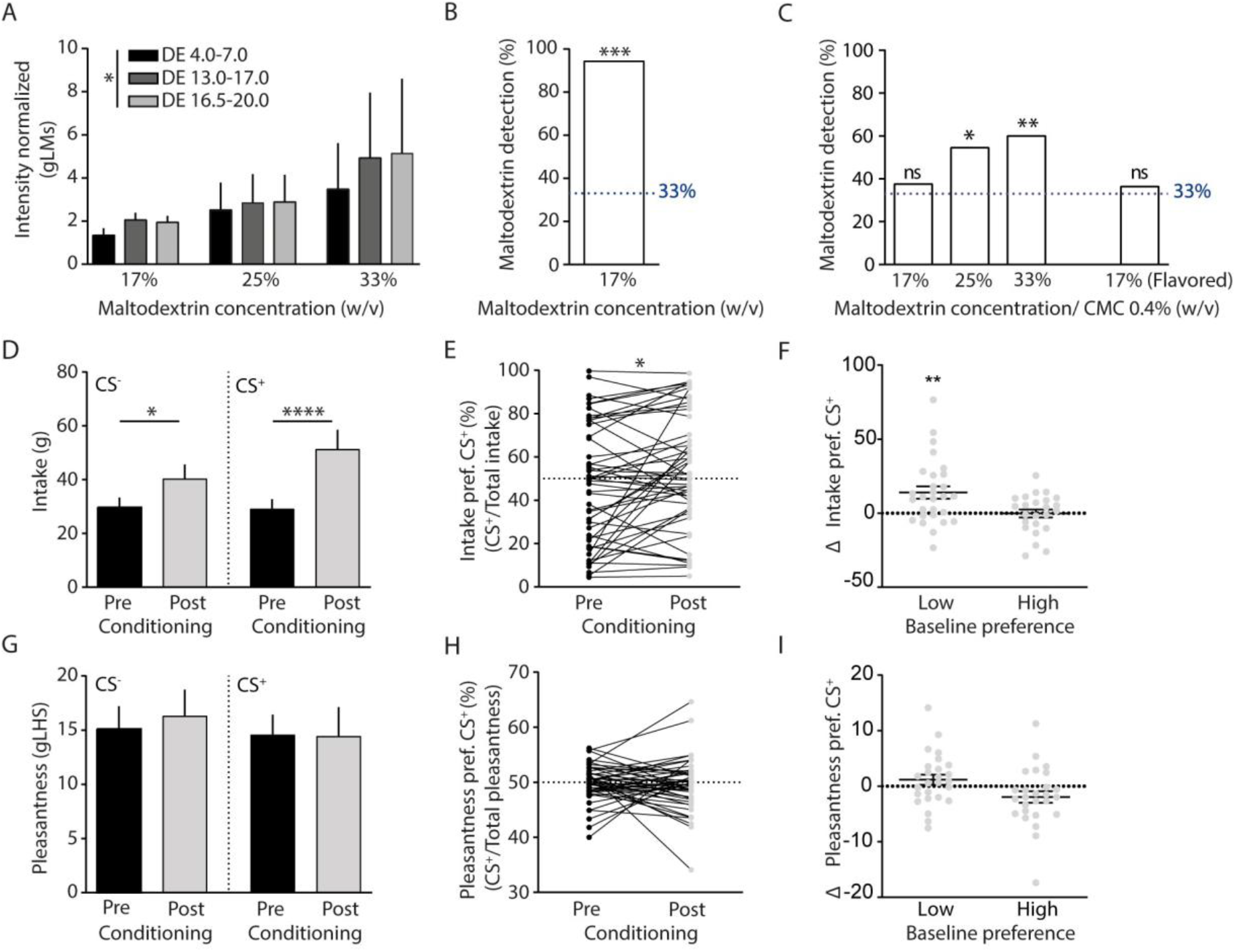
Development of a novel Flavour Nutrient Conditioning protocol. **(A)** Unflavoured maltodextrin yoghurt was tested at one of three different maltodextrin concentrations (17%, 25% and 33% w/v) in 3 different groups of healthy individuals. In each group participants consumed base yoghurt solution without maltodextrin and three solutions with maltodextrin at distinct dextrose equivalents (DE 4-7; DE 13-17 and DE 16.5-20). Each yoghurt solution was rated according to intensity and pleasantness. Intensity ratings of maltodextrin yoghurts, normalized to ratings of base yoghurt varied according to DE (F(2,72) =3.4, *P*=0.04), independently of the concentration tested, (F(2,36)=0.5, *P*=0.6; interaction: F(4,72) =0.7, *P*=0.6; mixed-model two-way ANOVA; n=39). **(B)** We conducted 3-alternative forced-choice tests, where one or two of three yogurt samples contained maltodextrin (DE4-7), and the other(s) were control yogurts. The percentage of participants that detected maltodextrin was significantly above the chance level of 33% (94.4%, p<0.0001; binominal test vs. 33%; n=18). **(C)** In similar 3-alternative forced-choice tests as in (B), but testing discrimination relative to carboxymethylcellulose (CMC, 0.4% w/v) rather than base yoghurt, the higher concentrations of maltodextrin (25% and 33%), were identified significantly above chance (54.5%, *P*=0.02, n=22; 60%, *P*=0.001, n=25; respectively), while 17% maltodextrin yoghurts with were not discriminated from CMC yoghurts (37.5%, *P*=0.22, n=23), as confirmed when flavoured yogurts were tested (36.4%, *P*=0.13, n=32). **(D)** After conditioning, participants significantly increased the intake of both CS+ and CS-flavours (time: F(1,51)=17.1, *P*=0.0001; stimulus: *F*(1,51)=1.2, *P*=0.3; post-hoc tests: CS^+^, *P*<0.0001, CS^-^*P*=0.02), with the interaction between the two factors suggesting differential effects between stimuli (F(1,51)=4.3, *P*=0.04; repeated measures two-way ANOVA; n=52). **(E)** Post-conditioning, preference for CS^+^, as measured according to intake, increased significantly, (t(51)=2.6, *P*=0.02, paired t-test, n=52). **(F)** Participants with low baseline intake preference for CS^+^ (<50%) had a significant post-conditioning, increase in preference for CS^+^ (t(27)=3.4, *P*=0.002, n=28), whereas those with high baseline preference (≥ 50%) maintained preference (t(23)=0.08, *P*=0.9, n=24, one sample t-test vs. 0). **(G)** We found similar pleasantness ratings both for CS^-^ and CS^+^ before and after conditioning, and unchanged preference for CS^+^, as measured by pleasantness **(H)**. **(I)** In calculations according to pleasantness ratings, we found no effect of conditioning on change of preference for CS^+^, irrespective of the baseline preference. In panels A, D, F, G and I, data is presented as mean ± standard error of the mean (SEM). gLMS/ gLHS - general labelled magnitude/hedonic scales. \**P* ≤ 0.05; \*\**P* ≤ 0.01; \*\*\*\**P* ≤ 0.0001.

### Flavour-nutrient conditioning occurs through increased intake, but not pleasantness ratings

In another group of healthy volunteers (n=52, **Table 1**), we then tested a FNC protocol using the optimized sweetened low-fat yogurt solutions, i.e., with either 17% DE 4.0-7.0 maltodextrin (+0.68Kcal/mL; unconditioned stimulus) or 0.4% CMC (+0.012 Kcal/mL; control stimulus), paired to two distinct flavours (0.3% w/v, respectively CS^+^ and CS^-^). During 2 days of home-conditioning with 150g CS^+^/Maltodextrin, alternated with 2 days of 150g CS^-^/CMC, no effects were found for day or stimulus on hunger, thirst, novelty, intensity, or pleasantness ratings (**Supplementary Figure 1 A, B, C, D and E)**. However, there was overall greater intake of CS^+^ than CS^-^ (F_(1,102)_=5.5, *P*=0.02), with less consumption in the 2^nd^ conditioning day (F_(1,102)_=8.1, *P*=0.005; interaction: F_(1,102)_=0.5, *P*=0.5; **Supplementary Figure 1 F**). Hunger, thirst and intensity ratings also did not differ significantly between pre-and post-conditioning days (**Supplementary Figure 1 G, H, and J)**, while novelty ratings decreased significantly after conditioning (F**_(_**_1,51)_=10.21, *P*=0.002), similarly for both stimuli (F_(1, 51)_=0.17, *P*=0.7; interaction: F_(1, 51)_=1.14, *P*=0.29; **Supplementary Figure 1 I)**, presumably as a result of multiple exposures during conditioning. Importantly, intake increased significantly in post-conditioning tests, relative to pre-conditioning (F_(1,51)_=17.1, *P*=0.0001), without significant differences between CS^-^ and CS^+^ (*F*_(1,51) =_ 1.2, *P*=0.3). However, there was a significant interaction between time and stimulus (F_(1,51)_=4.3, *P*=0.04; **Figure 1 D**), showing that this change was differential between stimuli. Indeed, the %preference for CS^+^, as calculated according to intake (intake %preference), significantly increased from pre to post-conditioning (t_(51)_=2.61, *P*=0.02; **Figure 1 E)**, suggesting that change in this measure (ΔCS^+^ preference = post-test minus pre-test CS^+^ %preference) may be used to assess efficacy of conditioning per individual. Indeed, while in participants with ≥50% CS^+^ intake %preference at baseline, ΔCS^+^ preference did not change significantly after conditioning (t_(23)_=0.08, *P*=0.9, n=24), a significant increase was found for those with low (< 50%) baseline intake %preference for CS^+^, suggesting that conditioning was restricted to this subgroup (t_(27)_=3.4, *P*=0.002, n=28; **Figure 1 F**; **Supplementary Tables 1** and 2). Pleasantness ratings, on the other hand, did not change significantly after conditioning (F_(1, 51)_=0.05, *P*=0.8), and were similar for CS^-^ and CS^+^ (F_(1,51)_=0.4, *P*=0.5; interaction: F_(1,51)_ = 0.2, *P*=0.8; **Figure 1 G)**. Consistently, CS^+^ %preference, when calculated according to pleasantness ratings (pleasantness %preference), did not differ between pre- and post-conditioning (*t*_51_=0.5, *P*=0.6; **Figure 1 H)**. Furthermore, in calculations according to pleasantness measurements, ΔCS^+^ preference did not differ from zero in those with low baseline pleasantness %preference for CS^+^ (t_(25)_=1.3, *P*=0.2, n=26), and had a close to significant reduction, rather than increase, in those with high baseline pleasantness %preference (t_(25)_=1.9, *P*=0.07, n=26; **Figure 1 I**). Importantly, sensitivity analysis excluding participants with BMI 25kg/m2 or greater revealed overlapping results to those obtained in the full sample (data not shown). Overall, these findings support that human flavour nutrient conditioning, when performed controlling for orosensory discrimination of maltodextrin, influences primarily implicit feeding decisions (i.e., increase in intake) rather than explicit assessments of stimuli (i.e., increase of pleasantness).

### Flavour-nutrient conditioning is conserved in obesity and after bariatric surgery

We then used the optimized FNC protocol to test a clinical cohort of patients from a bariatric surgery programme, where we recruited 34 eligible patients, 11 of whom with obesity approved for bariatric surgery and 23 after bariatric surgery (gastric bypass, n=13; sleeve gastrectomy, n=10), that were compared with a group of 27 healthy controls (**Supplementary Figure 2 A-B**). Groups differed significantly according to age (F_(2,60)_=12.7, *P*=0.00003), BMI (F_(2,60)_=106.3, *P*<0.00001) and years of formal education (F_(2,60)_=10.9, *P*=0.0001; **Table 1**), but were similar across most gustatory and psychometric variables except PFS scores (aggregate and food available; both *P=*0.04), addiction-like feeding behaviour (number of symptoms, *P*=0.004; diagnosis rate *P*=0.01) and external eating (*P*=0.001; **Table 1).**

Here, for FNC, flavours with lower baseline intake preference were selected to pair with maltodextrin, given that during FNC development, this was necessary for conditioning to be demonstrated, and reverse effects (i.e., reduction of intake preference) were not found when maltodextrin was paired with flavours with high baseline preference (please see **Figure 1 F**), as would be expected if effects resulted simply from regression to the mean^27^. Consequently, CMC was paired with the flavour with higher pre-conditioning intake preference, and maltodextrin paired with the flavour with lower preference. During conditioning, significant differences were not found for group nor for CS type regarding hunger, thirst, novelty and pleasantness ratings (**Supplementary Figures 3 A-C, E)**. However, intensity ratings varied according to CS type (F_(1, 50)_=5.2, *P*=0.03; **Supplementary Figure 3 D**), while intake varied according to group (F_(1, 50)_=7.5, *P*=0.001; interaction: F_(2, 50)_=1.0, *P*=0.4; **Supplementary Figure 3 F)**. Consistently with data from the original healthy volunteer group, while there were no effects for hunger, thirst and intensity, novelty ratings decreased from pre-to post-conditioning (*F*_(1,50)_=12.9, *P*=0.001; **Supplementary Figure 3 G-J**). Regarding the effects of conditioning on flavour preference, we found that ΔCS^+^ preference, as assessed by intake, increased significantly after conditioning (t_(52)_=3.6, *P*<0.001; n=53), with no significant differences significantly across healthy (n=24) obese (n=9) and surgical groups (n=20; F_(2, 50)_ = 1.9, *P*=0.2; **Figure 2 A)**. Regarding pleasantness ratings, ΔCS^+^ preference also did not vary significantly according to group (F_(2, 50)_ = 0.2, *P*=0.8; **Figure 2 B**) and, as observed in the initial experiments, did not reflect any significant effects of conditioning (t_(52)_=-1.4, *P*=0.2; n=53). In exploratory analyses, ΔCS^+^ preference did not differ between the sleeve and bypass groups, for neither intake (t_(18)_=1.1, *P*=0.3) nor pleasantness (t_(18)_=-0.2, *P*=0.9; n=20). Finally, sensitivity analyses excluding participants with BMI 25kg/m2 or greater from the healthy volunteer group revealed overlapping results to those obtained in the full sample (data not shown).

**Figure 2.**
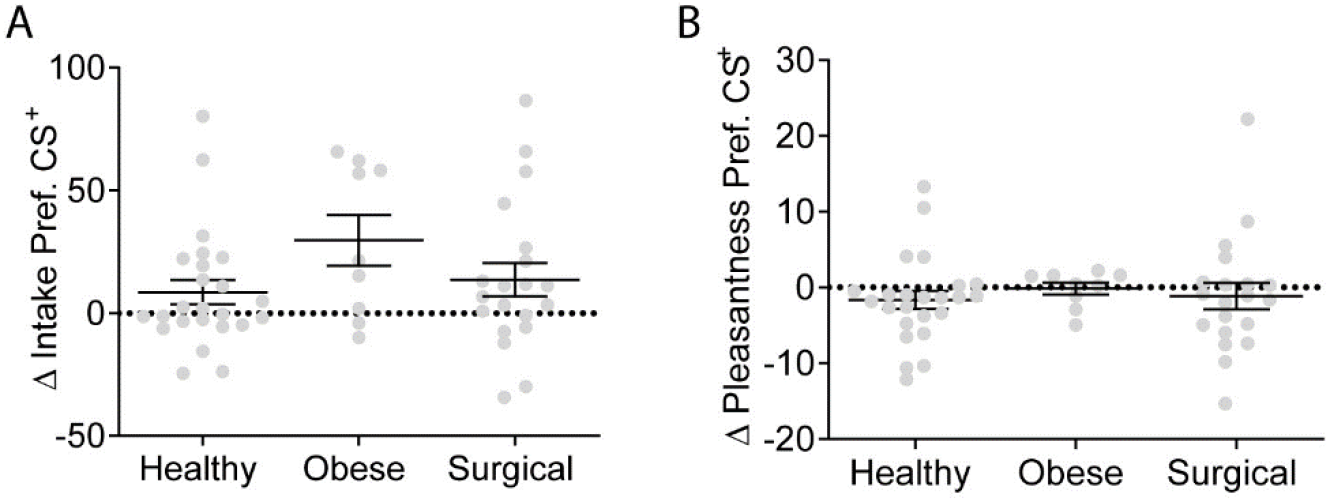
Measures of flavour-nutrient conditioning across the clinical study groups. **(A)** ΔCS^+^ preference as measured by intake, across the clinical study groups. A one-way ANOVA revealed a non-significant group effect (F(2, 50)=1.9, *P*=0.2), across healthy (n=24) obese (n=9) and surgical (n=20) groups. **(B)** ΔCS^+^ preference as measured by pleasantness ratings, across the clinical study groups. A one-way ANOVA revealed a non-significant group effect (F(2, 50) = 0.2, *P*=0.8), across groups. Data is presented as mean ± standard error of the mean (SEM). gLMS - general labelled magnitude scales. \**P* ≤ 0.05; \*\**P* ≤ 0.01.

### Reduced striatal DD2lR availability in obesity is recovered after bariatric surgery and may be related to postingestive conditioning after gastric bypass

While reduced availability of DD2lR is associated with extreme obesity^14, 28–30^, there is controversy relative to the possibility that bariatric surgery may normalize this effect^31–34^, with a need for further studies and some evidence of advantages in the use of [^123^I] IBZM SPECT^15^. We used this method to assess DD2lR availability in the groups of the clinical study and performed exploratory analyses to test associations with FNC in the surgical group. We found significant differences in DD2lR availability across study groups (F_(2,52)_=9.8, *P*=0.0002), that was lower in patients with obesity (n=11) when compared both with healthy volunteers (*P*=0.02, n=21) and surgical patients (*P*=0.0001, n=23), but did not differ between the surgical and healthy groups (*P*=0.2; **Figure 3 A and B**), with globally overlapping results in sensitivity analyses excluding participants with BMI 25kg/m2 or greater from the healthy volunteer group (data not shown). While we did not find differences in DD2lR availability between the gastric bypass (n=13) and sleeve gastrectomy (n=10) sub-groups (*P*=1.0), within the former group DD2lR availability was negatively correlated with ΔCS^+^ preference (intake) (r=-0.7, *P*=0.02, n=11; **Figure 3 C**), and positively correlated with DEBQ - restrained eating (r=0.8, *P*=0.01, n=11; **Figure 3 D**). Neither of these correlations were found in sleeve gastrectomy (**Figure 3 C and D**) or other groups (see Supplementary **Table 3** for details). Our results corroborate previous findings of decreased DD2lR availability in obesity and support the possibility that these obesity-related effects are reverted by bariatric surgery, similarly following gastric bypass and sleeve gastrectomy. Nevertheless, and in support of specificities of gastric bypass effects on postingestive conditioning, only after gastric bypass was there evidence of associations of DD2lR availability with FNC conditioning strength, as well as with restrained eating.

**Figure 3.**
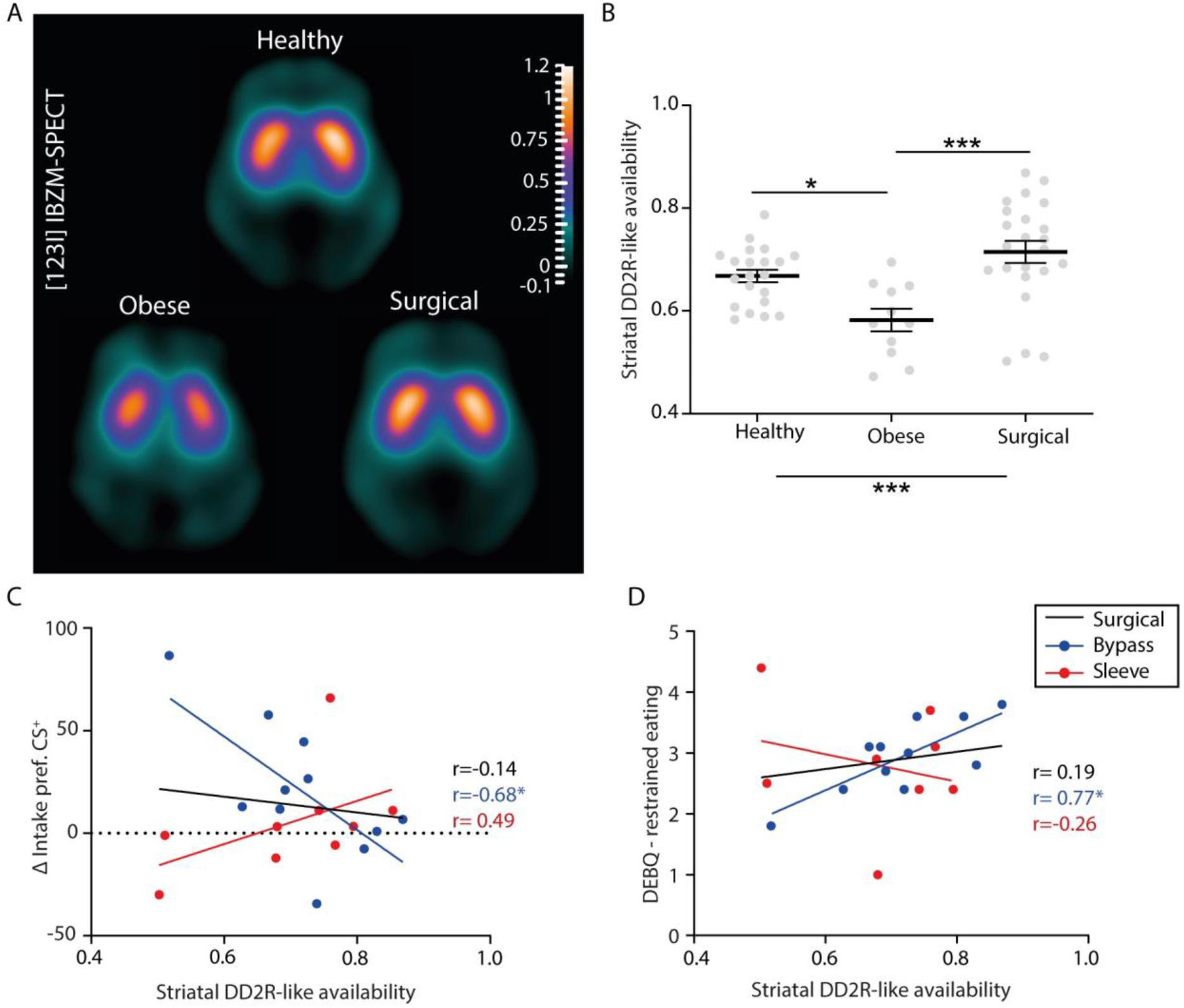
Striatal DD2lR availability in obesity and bariatric surgery and associations with postingestive conditioning and restrained eating. **(A)** Average [^123^I] IBZM images of the study groups. The figure shows the average group images in the striatal central transverse plane of healthy subjects (n=21; upper panel), patients with obesity (n=11; lower left panel), and surgical patients (n=23; lower right panel). **(B)**. A one-way ANOVA revealed a main group effect for striatal DD2lR availability across the clinical study groups (F(2, 52)=9.81, *P*=0.0002), with significant post-hoc Bonferroni tests supporting lower striatal BP for the obesity group relative to both surgical (*P*=0.0001) and healthy (*P*=0.02) groups. No differences were found between surgical and healthy groups (*P*=0.19). **(C)** Association between striatal DD2lR availability and ΔCS^+^ preference (intake) across surgery types (Surgical group, r=-0.14, *P*=0.6, n=20; Bypass, r=-0.68, *P*=0.02, n=11; Sleeve, r=0.49, *P*=0.18, n=9). **(D)** Association between Striatal DD2lR availability and the Dutch Eating Behaviour Questionnaire - restrained eating scores across surgery types (Surgical group, r=0.19, *P*=0.4, n=19; Bypass, r=0.77, *P*=0.01, n=11; Sleeve, r=-0.26, *P*=0.5, n=8). In panel B data is presented as mean ± standard error of the mean (SEM). *P ≤ 0.05; ***P ≤ 0.001

## Discussion

We showed that Flavour Nutrient Conditioning, when performed while controlling for explicit sensory effects of maltodextrin, is expressed primarily through implicit consumption decisions, rather than explicit assessments of flavour pleasantness ratings. Furthermore, this measure of postingestive learning was conserved across healthy volunteers, patients with severe obesity, and patients treated with bariatric surgery. Measures of DD2lR availability collected in the same clinical cohort were confirmed to be lower in patients with obesity than in healthy volunteers and patients after bariatric surgery, suggesting that obesity-related effects on DD2lR availability are reversible. Importantly, exploratory analyses showing, in the gastric bypass group only, associations between DD2lR availability and conditioning strength, as well as a measure of feeding behaviour regulation, suggest that processes of postingestive reinforcement may be of mechanistic relevance for this surgery type.

An important finding of our work is that the acquisition of preferences for flavours paired with calories from maltodextrin occurred according to food intake but not pleasantness ratings. Earlier results of FNC protocols with a similar conditioning period (4 days) and also using oral maltodextrin as a caloric source (112.5 Kcal) reported a small but significant increase in hedonic ratings for CS^+^ flavours^6, 7^. Those findings are consistent with a process whereby postingestive stimuli generate a hedonic value (i.e., ‘liking’ or induced sensation of pleasure)^2, 35^, interacting with other explicit components of food intake, such as flavour perception^2^. Those studies also had triangulation tests to address the possibility that individual participants were capable of detecting maltodextrin. However, we believe that the method used to define individual-level detection may have been insufficiently sensitive, since a similar approach used here, but across individuals, revealed clear signals for detection. Determining the conditions under which maltodextrin would not be detected was a fundamental step of our study, ensuring that the results of conditioning were determined primarily by postingestive stimulation. Indeed, when conducted under these conditions, FNC resulted in changes of intake of the flavour conditioned with maltodextrin, but not changes in hedonic ratings of pleasantness. Consistently, pre-clinical research supports the mediation of postingestive signals by striatal dopamine release^9, 11, 36^, that is thought to modulate food-seeking behaviours^11^ (i.e., ‘wanting’ or increased effort to obtain a reward), likely at an implicit level^2, 35^ as is supported here.

We confirmed results of preference change for CS^+^ flavours according to intake, as well as absent effects on pleasantness ratings, in a distinct cohort of patients from a bariatric surgery clinic and healthy controls. In addition, we found that postingestive conditioning was conserved across the several groups, without evidence for significant variation of the conditioning strength across groups. Since a chronic, excessive caloric intake marks obesity^1, 17^, it was plausible to hypothesize that patients with severe obesity would show altered conditioning strength relative to controls. Indeed, others have recently published data to support that brain responses to postingestive nutrient stimulation are attenuated or absent in patients with obesity, and suggested that these impairments may contribute to overeating^16^. Given the reconfiguration of the gastrointestinal tract induced by bariatric surgery^37^, and the resulting changes in food preferences^20–23^, we had also hypothesized that bariatric surgery would also impact postingestive reinforcement. Our results do not provide robust support for either of these hypotheses. We did not find deficits in conditioning strength associated to morbid obesity, nor changes resulting from bariatric surgery. Indeed a qualitative inspection of our results suggests that there may be enhanced, rather than impaired, postingestive conditioning in obesity, which is consistent with data in animal models, showing enhanced FNC in rats with diet-induced weight gain, relative both to rats that did not gain weight, and controls maintained on regular chow^38^. It is possible that our study lacked statistical power to address this question, since the obesity group was small, and there was significant variability in preference data. A larger sample of individuals with obesity, and optimized procedures to study postingestive conditioning, may be needed for further research these questions.

Previous findings of lower DD2lR availability in patients with obesity when compared with healthy individuals, described with similar methods as those used here^39^ or with [^11^C]raclopride positron emission tomography (PET)^40^, were confirmed, as well as the association of BMI and DD2lR availability among patients with morbid obesity^15, 40^. Our findings of higher DD2lR availability in the surgical group are globally consistent with a previous study using [^123^I]IBZM SPECT and describing a significant increase in DD2lR availability in women with morbid obesity two years after gastric bypass, but still with reduced levels in comparisons with age-matched controls^33^. We found a complete reversal to levels similar to those in healthy volunteers on average 2.5 years after surgery, that may have been due to study design and/or to a greater diversity in our sample. Indeed, we did not restrict recruitment only to women, and studied patients treated with gastric bypass as well as sleeve gastrectomy groups. While DD2lR availability did not differ between surgery types, it is possible that this may have contributed to small differences relative to published data. DD2lR availability, as assessed here with [^123^I]IBZM SPECT, is a static representation of dopaminergic physiology, and the changes associated with obesity may thus reflect decreased expression of the receptor and/or greater occupancy by dopamine^14^. van Galen et al^16^ described impairments of the striatal dopamine response to intragastric lipids, but not glucose, in patients with obesity which, albeit the absence of direct comparisons with healthy volunteers, is not suggestive of enhanced dopamine release^16^. In animal research, downregulation of dopamine D_2_ receptors was shown to result from consumption of energy-dense diets^41, 42^, but data on the impact of obesity on dopamine responses to food is lacking. Additional research is needed to fully understand the association between obesity, weight-loss and dopamine homeostasis.

Importantly, in exploratory analyses, we found a moderate to strong inverse association between DD2lR availability and postingestive conditioning strength, in the gastric bypass group only. In the same surgical group, DD2lR availability had a strong direct association with restrained eating that, in turn, had a moderate to strong negative correlation with conditioning strength. Volkow N et al.^43^ showed that high-restrained eaters had more significant striatal dopamine responses to food stimulation, as assessed with [^11^C]raclopride PET, with higher restraint suggested to reflect a preventive adaptation strategy to minimize exposure to salient food cues^57^. While, to our knowledge, there are no studies addressing the effects of gastric bypass on striatal dopamine responses, our results suggest that, after gastric bypass, patients with the largest increase of DD2lR availability are also those with greater use of restraint as a coping behaviour and with the least sensitivity to postingestive conditioning. These associations were absent for sleeve gastrectomy, where non-significant correlations in the opposite direction were found. Other studies have described differential effects of gastric bypass and sleeve gastrectomy on food reward-related measures^21^. It is possible that distinct methods for bariatric surgery, as well as variations within the same surgery type, may alter the vagal mediation of postingestive signals^11^, but evidence to support importance of the vagus nerve for weight loss and appetite suppression after gastric bypass, collected in rodents, is mixed^44, 45^. Further research, specifically designed to test the importance of postingestive reinforcement in the context of gastric bypass, is needed to address the hypotheses raised here.

### Limitations

The results of this study should be interpreted according to its limitations. Our FNC protocol is limited by the fact that the four conditioning days were conducted at home, to avoid loss to follow-up, but also limiting experimental control over this phase of the experiment. Despite our efforts to minimize these limitations, namely the use of saliva samples to increase compliance for fasting and exclusion of participants with low adherence to conditioning procedures, there is a possibility for self-report bias, that we cannot account for. On the other hand, we did not *a priori* match groups in the clinical study for age, gender, or education. A strict matched case-control design in the clinical study would have been an asset, but is hindered due to the challenges in recruitment within the clinical groups, particularly for a complex protocol as described here. Furthermore, the obesity and surgical groups were not assessed prospectively, in an attempt to avoid the effects of learning on repeated exposures to the FNC paradigm, and also due to the known challenges of longitudinal follow-up in this clinical population^46^. Challenges in recruitment of the clinical population was also reflected in small sample sizes, that may have limited statistical power. Larger studies addressing more restricted hypotheses (e.g., differences in obesity vs. controls or the impact of gastric bypass) are needed to replicate and/or expand these results. Finally, we assessed DD2lR availability in a static protocol, after the FNC protocol. Ideally, future research should quantify brain responses to postingestive conditioning in real-time.

### Conclusions

Using a novel method for FNC in humans, we showed that postingestive reinforcement occurs in healthy subjects and is expressed in implicit behaviour rather than explicit pleasantness scores. Furthermore, this postingestive learning was conserved in patients with obesity and post-bariatric patients, suggesting that it may play a role in feeding behaviour regulation across these groups. However, reduced DD2lR availability was found in patients with obesity when compared to post-bariatric patients, as well as healthy volunteers, with associations between this variable and postingestive conditioning strength, specifically for patients treated with gastric bypass. Thus, while postingestive reinforcement is conserved in obesity and after bariatric surgery, it may play a role in the impact of gastric bypass on feeding behaviour regulation, deserving further research the address this hypothesis.

## Methods

### Study design and participants

Healthy volunteers were recruited from the community, initially to optimize conditions for use of maltodextrin in FNC (‘Maltodextrin optimization’ group), and then to test and optimize a controlled protocol for FNC (’FNC development’ group). Inclusion criteria were age between 18 and 65 years and general good health as determined by the investigator. Exclusion criteria, assessed at entry into the study, were active acute respiratory infection, active neurological or psychiatric disease, active gastrointestinal, hepatic, or pancreatic disease, diabetes, illicit substance use or alcohol abuse; use of any neuropsychiatric medication (including anxiolytics, antipsychotics, antidepressants, anticonvulsants, stimulants, anti-dementia medication, dopamine agonists and opioid analgesics) or antidiabetic medication (including glp-1 agonists); illiteracy, or otherwise not understanding instructions for the study; prior major gastrointestinal surgery and/or intra-gastric balloon in the previous 12 months; history of food allergies; pregnancy or breastfeeding. The clinical cohort consisted of consecutive patients at a tertiary care outpatient centre specialized in the surgical treatment of obesity, belonging to Centro Hospitalar de Lisboa Ocidental, E.P.E., in Lisbon, Portugal. The cohort included a group of patients approved for bariatric surgery and on the waiting list (obese group) and a group of patients that had received bariatric surgery (surgical group). The latter were recruited no less than 1.5 and no more than 4 years after either gastric bypass or sleeve gastrectomy, at a time when patients are expected to be weight stable and capable of consuming small volumes of liquid. Approval for bariatric surgery followed standard criteria as defined by the Portuguese National Health Service^24^. Exclusion criteria for patients were equivalent to those mentioned above, except for BMI, and prior major gastrointestinal surgery for the surgical group only. Patients were identified by the clinical team, and those consenting to be contacted were screened by phone call. Those not excluded were further assessed for eligibility at admission into study. For patients, we retrieved the surgery date and type from clinical files to avoid self-report bias. An additional group of healthy volunteers were recruited for comparisons with patients. Exclusion criteria were equivalent to those mentioned above as well as obesity (BMI ≥ 30 Kg/m^2^) and underweight (BMI < 18.5 Kg/m^2^). Ethics Committees approved the study protocol at Champalimaud Foundation, Lisbon Academic Medical Centre, and Centro Hospitalar Lisboa Ocidental. Informed consent was obtained in writing from each participant. The possibility of discontinuing participation at any time during the study was given to all participants. All data were de-identified.

### Solutions

Yoghurt (34 Kcal, 0.1 g of fat, 4.3 g of carbohydrates and 4.0g of protein per 100g) was purchased from a national commercial provider (Continente, Portugal) and were stored at 4°C. Sucralose, maltodextrin and carboxymetilcellullose (Sigma Aldrich) and flavours (Nature’s Flavours, Orange, CA, USA) were stored at room temperature. Milli-Q® water was obtained from our institutional distilled water system. All yoghurt-based solutions were prepared daily under sterile conditions, and stored at 4°C until 1 h prior to each experiment, when they were transferred to room temperature. Maltodextrin was first diluted in 1/3 of the intended final yoghurt solution volume in Milli-Q® water. The solution was dissolved using a plate heater and a magnetic stirrer, at a 90°C (Ohaus, USA). Once this solution was again at room temperature, the final intended volume was completed with 2/3 yoghurt to obtain final maltodextrin yoghurt at concentrations of 17%, 25% or 33% (w/v; respectively 0.68Kcal/mL, 1Kcal/mL, 1.32Kcal/mL). CMC (carboxymethylcellulose) yoghurt solutions were similarly prepared to obtain a final concentration of 0.4% (w/v; (0.012 Kcal/mL). A base yoghurt solution was prepared with 1/3 Milli-Q® water and 2/3 yoghurt (v/v). All yoghurt solutions had sucralose added at a concentration of 0.01% (w/v) and, for flavoured solutions (cashew, lychee, tamarind, cider, black currant, and pomegranate), the selected flavour was added at a concentration of 0.3% (w/v).

### Optimizing maltodextrin concentration and dextrose equivalents

In a first cohort of healthy volunteers, psychophysical assessments of distinct maltodextrin concentrations and dextrose equivalents (DE) were performed. Participants were divided into 3 groups, with each group testing one concentration of maltodextrin (17%, 25% or 33%). For each concentration, in addition to a base yoghurt solution, participants sampled 3 solutions of maltodextrin enriched yoghurt, all at the same concentration but prepared with maltodextrin of distinct dextrose equivalents (DE), namely DE 4-7, DE 13-17 and DE 16.5-20. The 4 yoghurt solutions were presented in randomized order, and immediately rated according to intensity (0 to 100 mm Visual Analogue Scale - VAS^47^) and pleasantness (−100 to 100 mm, general Labelled Hedonic Scale - gLHS^48^).

### Discrimination tests to assess maltodextrin detection

Additional groups of healthy volunteers performed a 3-alternative forced-choice (3-AFC) tests to assess discriminability of sweetened maltodextrin yoghurt solutions. In an initial test, discrimination of 17% DE 4-7 maltodextrin was tested against a base yoghurt solution. The 3-AFC test consisted in presentation of one or two cups with 17% maltodextrin yoghurt solution, with the remaining (respectively two or one) cup(s) containing the base yoghurt solution. Participants were asked to sample all three of the yoghurts, and then select the one that was different from the other two. In three additional cohorts of healthy volunteers, each of the three different maltodextrin yoghurt concentrations (17%, 25% or 33%), all prepared with DE 4-7 maltodextrin, were contrasted in 3-AFC tests with CMC yoghurt solution (0.4%). In a final group of healthy participants, the contrast between 17% maltodextrin and 0.4% CMC yoghurt solutions was repeated, but with one of six flavours (cashew, lychee, tamarind, cider, black currant, and pomegranate; 0.3%) added to the solutions used in each 3-AFC test.

### Flavour-Nutrient Conditioning

Experimental sessions occurred on six consecutive days, all of which following an overnight fast of 8-10h. Participants attended the laboratory on the first (pre-conditioning) and last (post-conditioning) days, with four conditioning days performed at home between the first and last test days. During the experiments, Milli-Q® water at room temperature was available for consumption if desired by the participant. On the first day, participants were assessed for height and weight with a digital scale and a mechanical stadiometer (Kern & Sohn GmbH, Balingen, Germany) with light clothes and without shoes. At the end of that day each participant completed a gustatory psychophysics (taste strips method for citric acid, sodium chloride, sucrose and quinine hydrochloride; taste thresholds assessed with electrogustometry^49^) and psychometric evaluation of reward-related feeding behaviour (Power of Food Scale^13, 50–52^, Yale Food Addiction Scale^53, 54^, Dutch Eating Behaviour Questionnaire^55, 56^ and Food Action Rating Scale^57^), as described previously (please see Ribeiro et al 2021^24^ for details). In the pre-conditioning day, after collecting ratings of hunger and thirst on 0 to 100 mm Visual Analogue Scales (VAS), participants were presented with samples of six differently flavoured (cashew, lychee, tamarind, cider, black currant, and pomegranate) CMC yoghurt solutions, presented in random order for ratings of stimulus novelty (VAS), intensity (0 to 100 mm gLMS) and pleasantness (−100 to 100 mm gLHS). We selected two flavoured beverages for each participant, based on high novelty and similar moderate pleasantness. Each subject then performed 6 trials of 3-AFC discrimination tests contrasting the two selected flavours, with a revision of the flavours selected if correct discrimination was not obtained in at least 4 of the trials. Participants were excluded if two flavours with pleasantness rated above 0 in the gLHS and that were correctly discriminated could not be identified. We then presented the two flavoured CMC yoghurt solution in two large white cups for ad libitum consumption/intake, and measured weight (g) before and after consumption (intake measurement). In the initial cohort of healthy volunteers, one of these flavours was randomly chosen to pair with maltodextrin during home conditioning (CS^+^, +102 Kcal), while the alternate flavour was paired with CMC (CS^-^, +1.8 Kcal). For the clinical experiment maltodextrin was always paired with the least preferred flavour, as assessed according to intake during ad libitum consumption. In the following four conditioning days, at home, subjects were instructed to maintain overnight fasting, after which they should consume yoghurt solutions offered to them in the pre-conditioning day in sterilized glass bottles, containing 150g of yoghurt, and that they were instructed to conserve at 4°C. Bottles were labelled with the consumption date and a letter code assigned to CS^+^ or CS^-^ flavours, so that they consumed either a 17% maltodextrin yoghurt solution, paired with the CS^+^ flavour, in two non-consecutive days, or a 0.4% CMC yoghurt solution, paired with the alternate flavour (CS^-^), in the two alternate days, with the order of flavours randomized. Participants were asked to return any yoghurt that they did not consume for quantification of intake (g), with exclusion from analysis if mean home consumption was, on average, less than 25g in total or for any of the flavours. They were also asked to perform ratings of hunger and thirst prior to consumption, and then of stimulus novelty, intensity and pleasantness. In the post-conditioning day, we presented the same six-flavour sequence of CMC yoghurt solutions, as on the first day, for the same process of flavour rating. Then, the two flavours selected for conditioning were given for ad libitum consumption. The main outcomes of this protocol were changes in %preference for CS^+^, assessed according to intake (CS^+^ intake %preference) or pleasantness ratings (CS^+^ pleasantness %preference) from pre-to post-conditioning. In participants of the clinical study, at the end of the last day, single-photon emission computed tomography (SPECT) scans were performed.

### Striatal DD2lR availability imaging

We assessed striatal DD2lR availability using SPECT with [^123^I]-Iodobenzamide ([^123^I]IBZM, GE Healthcare, Eindhoven, NL). Participants were scanned early in the afternoon for approximately 30 minutes, 2 hours after a bolus injection of 185 MBq of [^123^I]IBZM. Each participant was pre-treated with potassium iodide to block thyroid uptake of free radioactive iodine (^123^I). SPECT imaging was performed using a Philips BrightView gamma camera (Philips Healthcare, Eindhoven, NL), equipped with low energy and high-resolution collimators. Image reconstruction was performed using the Astonish algorithm (Philips Healthcare, Eindhoven, NL), an optimized 3-dimensional ordered subsets expectation maximization (3D-OSEM) algorithm. After reconstruction, images were corrected for attenuation using the Chang method (linear attenuation coefficient of 0.11 cm^-1^) and the Hanning filter (cut-off 1.0). SPECT images were reconstructed with cubic voxels of 4.664 mm width and a region of interest (ROI) analysis was performed for quantification based on an automated software validated for brain [^123^I]FP-CIT SPECT scans^58^. This software was adapted for brain [^123^I]IBZM and validated against semi-automated quantification performed by experienced nuclear medicine physicians (data not shown). The primary outcome of ROI analysis was the striatal binding potential (BP) in the ROIs, which is a proxy for striatal DD2lR (i.e., D2 and D3 receptors) binding. When the radiopharmaceutical reaches equilibrium, the BP can be obtained as the ratio of the specific uptake and the non-specific uptake, as follows:

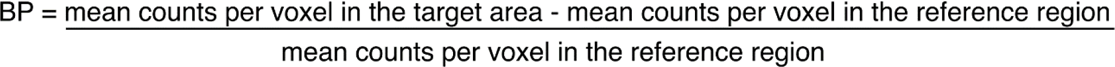

The reference region was a portion of the occipital lobe where D2 and D3 receptors are absent. The software quantifies the BP in six striatal ROIs (right caudate, right putamen, right striatum, left caudate, left putamen, and left striatum). For statistical analyses, the left and right striatum mean values were considered.

### Data analysis

Analyses were performed using SPSS version 29 (SPSS Inc., Chicago, IL, USA). Graphs were produced in GraphPad Prism version 8.0 (GraphPad Software, La Jolla, CA, USA) and edited in Adobe Illustrator version 2022 (Adobe Inc., San Jose, CA, USA). Data for continuous measurements is presented as mean ± standard error of mean (SEM). Assessment of normal distribution of continuous measurements was performed according to visual inspection of distribution as well as analysis of kurtosis, skewness and comparison between mean and median. Demographic, clinical, psychometric and psychophysical data was compared between groups using independent samples t-tests or one-way analysis of variance (ANOVA) for continuous variables and χ2 tests for categorical variables. In the maltodextrin optimization group, normalized intensity ratings were analysed using mixed model two-way ANOVA according to concentration (between subjects) and maltodextrin DE (within subjects). Proportion of participants correctly discriminating maltodextrin yoghurts in 3-AFC tests was contrasted to 1/3 (chance level) using binomial tests. To determine changes in raw intake and pleasantness ratings from pre-to post-conditioning in the healthy group (’FNC development’), we used repeated-measures two-way ANOVA with intake (g) or pleasantness ratings (mm) as independent variables, according to a time factor pairing pre-to post-conditioning days (*pre-post*) and a stimulus factor comprising CS^-^ or CS^+^ flavours (CS*^-^ vs* CS*^+^*). Intake %preference for CS^+^ was calculated as [CS^+^ intake/ (CS^-^ + CS^+^ intake) *100]. The same formula, but using gLHS pleasantness ratings, was used to calculate pleasantness %preference for CS^+^. In this case, we transformed pleasantness ratings by adding the amount needed for the minimum value to be 1 (’+101’). To determine changes in preference for CS^+,^ we used paired t-tests to compare pre-to post preferences according to intake or pleasantness. The difference between the pre- and post-conditioning preference for CS^+^ (Δpreference CS^+^), calculated according to either intake or pleasantness, was analysed separately according to respective baseline %preference, namely low (<50%) and high (≥ 50%), using one-sample t-tests contrasting against zero, to test whether there were significant changes in preference in each group. To compare groups in the clinical cohort for Δpreference CS^+^ (intake) or for DD2lR availability, we used a one-way ANOVA. Other data from FNC experiments either comparing pre-vs. post-conditioning days (hunger, thirst, novelty, intensity), or data from home-conditioning (hunger, thirst, novelty, intensity, pleasantness, intake) was analysed using repeated-measures two-way ANOVA, paired t-tests or mixed-model two-way ANOVA in the case of between-group analyses. Across ANOVA analyses, Bonferroni post-hoc tests were performed as planned. Exploratory associations between DD2lR BP, Δ intake preference for CS^+^, gustatory and psychometric feeding behaviour variables were determined using Pearson’s correlation (r). A two-tailed p-value of 0.05 was selected as the significance level for all analyses.

## Author contributions

GR, ABF, DCC, RMC and AJOM conceived and designed the experiments; GR and ABF performed experiments in healthy volunteers; GR, ABF, JSD, MO, and CL enrolled patients and acquired data for the clinical experiment; GR, ABF, FPMO, DCC and AJOM, analysed the data; GR, ABF and AJOM wrote the manuscript, that was critically reviewed by all other authors.

## Data Availability

All data produced in the present study are available upon reasonable request to the authors.

## Acknowledgements

GR was funded by doctoral fellowships from Universidade de Lisboa (BD/2015Call) and Fundação para a Ciência e Tecnologia (FCT; SFRH/BD/128783/2017). GR and AJOM were supported by the Champalimaud Foundation through a Clinical Kickstarter grant. ABF was supported by a postdoctoral fellowship (SFRH/BPD/880972/2012) and is supported by a postdoctoral contract (DL 57/2016/CP1483/CT0001) and grant (PTDC/SAU-NUT/3507/2021) from FCT. AJOM was supported by grants from the BIAL Foundation (176/10) and from (FCT), through a Junior Research and Career Development Award from the Harvard Medical Portugal Program (HMSP/ICJ/0020/2011), and is funded by a Starting Grant from the European Research Council (ERC) under the European Union’s Horizon 2020 research and innovation programme (grant agreement No. 950357).

## Notes

### Competing Interest Statement

According to ICMJE guidelines for Disclosure of Potential Conflicts of Interest, none were reported by GR, ABF, FPMO, JSD, MO, CL, DCC and RMC. AJO-M was national coordinator for Portugal of a non-interventional study (EDMS-ERI-143085581, 4.0) to characterize a Treatment-Resistant Depression Cohort in Europe, sponsored by Janssen-Cilag, Ltd (2019-2020), national coordinator for Portugal of trials of psilocybin therapy for treatment-resistant depression, sponsored by Compass Pathways, Ltd (EudraCT number 2017-003288-36), and of esketamine for treatment-resistant depression, sponsored by Janssen-Cilag, Ltd (EudraCT NUMBER: 2019-002992-33); is recipient of a grant from Schuhfried GmBH for norming and validation of cognitive tests; received payment, honoraria or other support from Janssen, Angelini, MSD, Neurolite AG, and the European Monitoring Centre for Drugs and Drug Addiction; is Vice-President of the Portuguese Society for Psychiatry and Mental Health, and Head of the Psychiatry Working Group for the National Board of Medical Examination (GPNA) at the Portuguese Medical Association and Portuguese Ministry of Health.

### Clinical Trial

ISRCTN17965026

### Funding Statement

Gabriela RIbeiro was funded by doctoral fellowships from Universidade de Lisboa (BD/2015Call) and Fundacao para a Ciencia e Tecnologia (FCT; SFRH/BD/128783/2017). Gabriela Ribeiro and Albino J.Oliveira-Maia were supported by the Champalimaud Foundation through a Clinical Kickstarter grant. Ana B. Fernandes was supported by a postdoctoral fellowship (SFRH/BPD/880972/2012) and is supported by a postdoctoral contract (DL 57/2016/CP1483/CT0001) and grant (PTDC/SAU-NUT/3507/2021) from FCT. Albino J. Oliveira Maia was supported by grants from the BIAL Foundation (176/10) and from (FCT), through a Junior Research and Career Development Award from the Harvard Medical Portugal Program (HMSP/ICJ/0020/2011), and is funded by a Starting Grant from the European Research Council (ERC) under the European Union's Horizon 2020 research and innovation programme (grant agreement No. 950357).

### Author Declarations

Ethics Committee of Champalimaud Foundation, Champalimaud Foundation, Lisbon Academic Medical Centre, and Centro Hospitalar Lisboa Ocidental gave ethical approval for this work.

