## Supplemental Material for "Postingestive reinforcement is conserved in obesity and after bariatric surgery"

#### Supplemental figures

**Supplementary Figure 1 -** Complementary conditioning measures in healthy subjects.

**Supplementary Figure 2 -** Flow diagrams of the clinical study groups.

**Supplementary Figure 3 -** Complementary conditioning measures across the clinical study groups.

#### Supplemental Tables

**Supplementary Table 1 -** Demographic characteristics of healthy subjects.

**Supplementary Table 2 -** Gustatory and psychometric measures of feeding behaviour in healthy subjects.

**Supplementary Table 3 -** Associations between striatal dopamine DD2lR availability, the conditioning strength, and feeding behaviour.

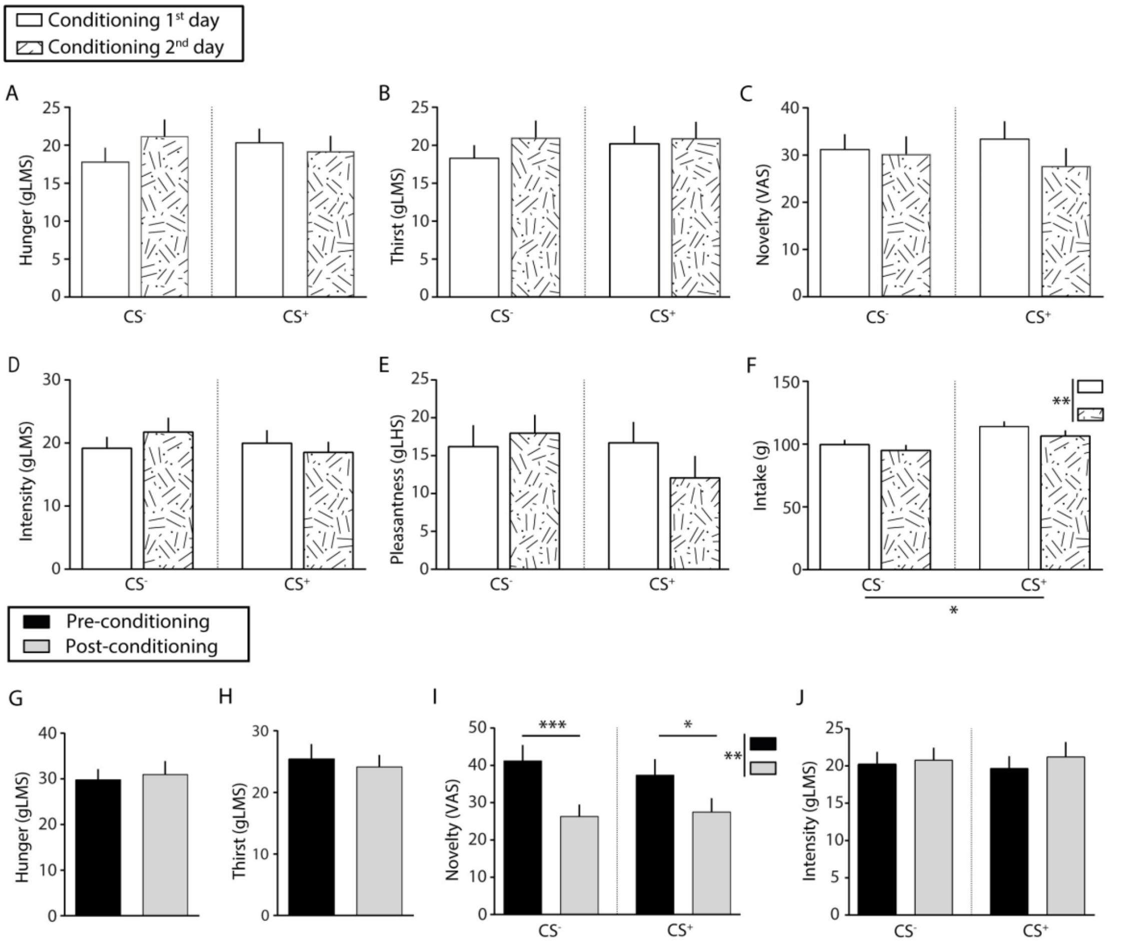

**Supplementary Figure 1. Complementary conditioning measures in healthy subjects.**

Across conditioning for $CS^-$ and $CS^+$ flavors, ratings did not vary according to stimulus or conditioning day for (**A**) **Hunger** (Stimulus: $F_{(1,102)}$=0.04, *P*=0.8; Day: $F_{(1, 102)}$=3.0, *P*=0.08; Interaction: $F_{(1,102)}$=9.9, P=0.9) **(B) Thirst** (Stimulus: $F_{(1,102)}$=0.1, p=0.8; Day: $F_{(1, 102)}$=3.2, *P*=0.07; Interaction: $F_{(1,102)}$=0.08, P=0.8) (**C**) **Novelty** (Stimulus: $F_{(1,102)}$=0.002, *P*=0.9; Day: $F_{(1, 102)}$=2.7, *P*=0.1; Interaction: $F_{(1,102)}$=1.3, *P*=0.3) (**D**) **Intensity** (Stimulus: $F_{(1,99)}$=0.3, *P*=0.6; Day: $F_{(1, 99)}$=0.09, *P*=0.8; Interaction: $F_{(1,99)}$=1.2, *P*=0.3) and (**E**) **Pleasantness** (Stimulus: $F_{(1,98)}$=0.9, *P*=0.3; Day: $F_{(1,,98))}$=0.4, *P*=0.5; Interaction: $F_{(1,98)}$=1.9, *P*=0.2). **(F) Intake** volumes were higher for $CS^+$ than $CS^-$ ($F_{(1,102)}$=5.5, *P*=0.02), and decreased across conditioning days ($F_{(1,102)}$=8.1, *P*=0.005; Interaction: $F_{(1,102)}$=0.5, *P*=0.5; repeated-measures 2-way ANOVA).
**(G) Hunger** ratings remained stable from pre-to post conditioning ($t_{(50)}$= 0.3, *P*=0.8) as well as **(H) Thirst** ratings ($t_{(50)}$= 0.5, *P*=0.6; paired t-test). **(I) Novelty** ratings significantly decreased from pre to post-conditioning ($F_{(1,51)}$=10.2, *P*=0.002; post-hoc $CS^-$, *P*=0.0001; post-hoc $CS^+$, *P*=0.01) but similarly for both stimuli ($F_{(1, 51)}$=0.17, *P*=0.7; Interaction: $F_{(1, 51)}$=1.1, *P*=0.3). **(J) Intensity** ratings remained similar from pre to post-conditioning ($F_{(1,51)}$=0.6, *P*=0.6), for both $CS^-$ and $CS^+$ flavors ($F_{(1,51)}$=0.0003, *P*=0.9; Interaction: $F_{(1,51)}$=0.2, P=0.7; repeated-measures 2-way ANOVA).

Bar graphs represent mean ± standard error of the mean (SEM).
gLMS/ gLHS - general labeled magnitude/hedonic scale.
VAS: Visual Analogue Scale.
$^*P \leq 0.05$; $^{**}P \leq 0.01$; $^{***}P \leq 0.001$

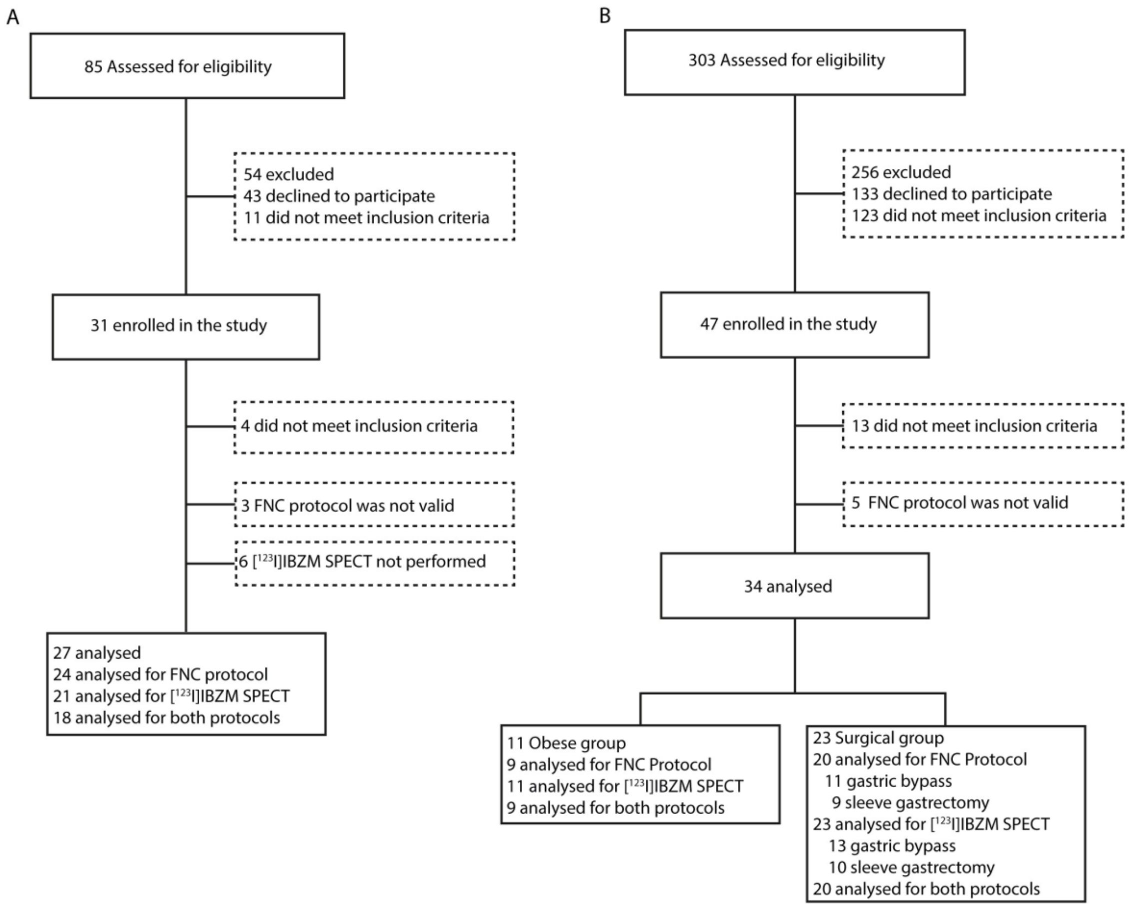

**Supplementary Figure 2. Flow diagrams of the clinical study groups.**

**(A)** Flow diagram of recruitment of the healthy control group. Six volunteers did not perform SPECT due to failures in $[^{123}I]$IBZM delivery or malfunction of the gamma camera. In 3 participants there was an error in FNC protocol.

**(B)** Flow diagram of the recruitment of the obesity and surgical groups. Across both groups, 5 participants were excluded from analysis of FNC according to exclusion criteria mentioned in methods.

FNC: Flavour Nutrient Conditioning protocol; $[^{123}I]$IBZM SPECT: $[^{123}I]$ iodobenzamide ($[^{123}I]$IBZM) single photon emission computed tomography (SPECT).

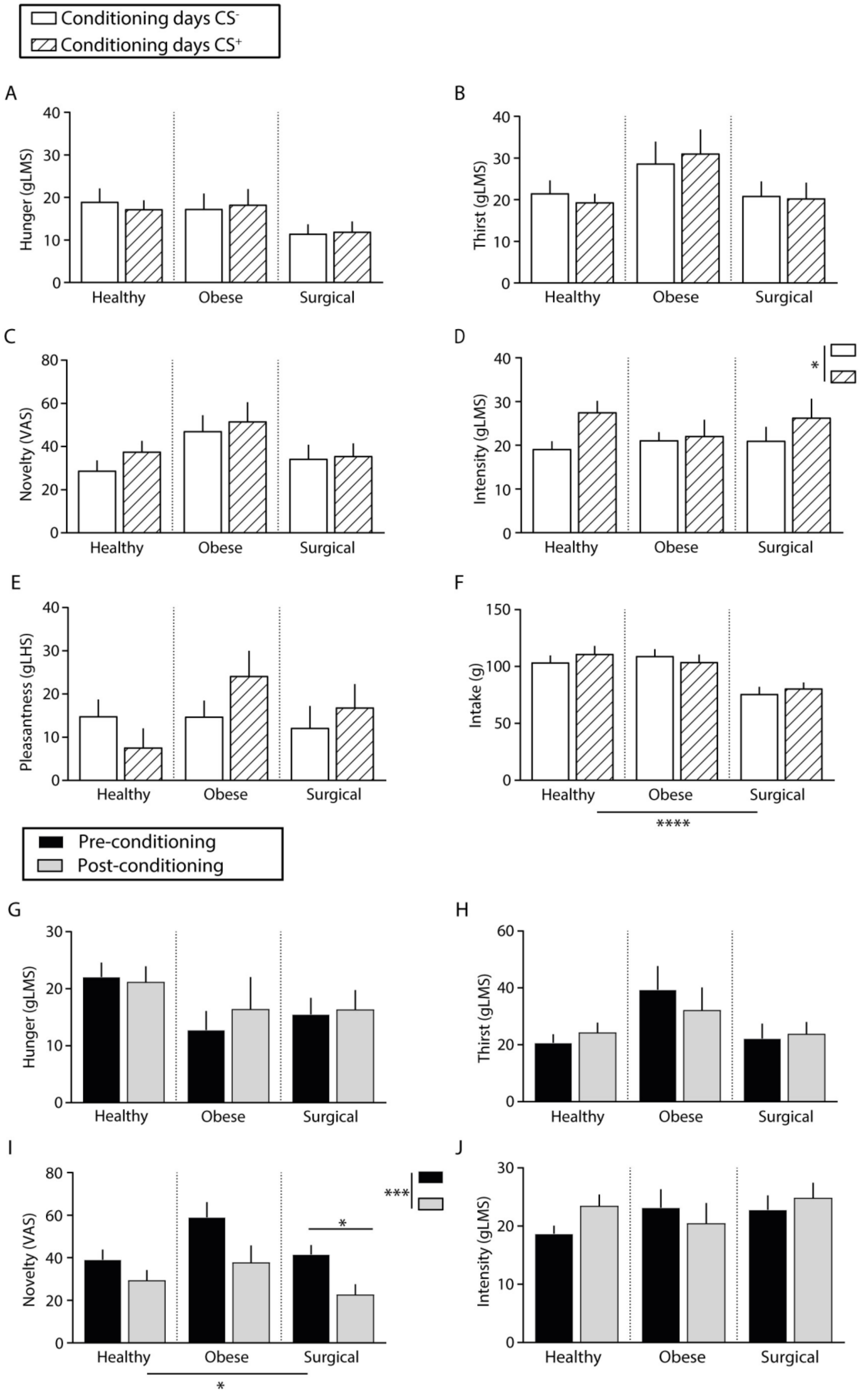

Conditioning days CS⁻
Conditioning days CS⁺
A
Hunger (gLMS)
Healthy
Obese
Surgical
B
Thirst (gLMS)
C
Novelty (VAS)
D
Intensity (gLMS)
*
E
Pleasantness (gLHS)
F
Intake (g)
****
Pre-conditioning
Post-conditioning
G
Hunger (gLMS)
H
Thirst (gLMS)
I
Novelty (VAS)
***
*
J
Intensity (gLMS)

### Supplementary Figure 3. Complementary conditioning measures across the clinical study groups.

Across conditioning for $CS^-$ and $CS^+$ flavours, ratings did not vary according to stimulus or group for **(A) Hunger** (Stimulus: $F_{(1, 50}$=0.003, *P*=0.96; Group: $F_{(2, 50)}$=2.2, *P*=0.1; Interaction: $F_{(2, 50)}$= 0.1, *P*=0.7); **(B) Thirst** (Stimulus: $F_{(1, 50)}$=0.01, *P*=0.9; Group: $F_{(2, 50)}$=1.5, *P*=0.22; Interaction: F $_{(2, 50)}$= 0.96, *P*=0.4 and **(C) Novelty** (Stimulus: $F_{(1, 50)}$=1.6, *P*=0.2; Group: $F_{(2, 50)}$=1.8, *P*=0.17; Interaction: $F_{(2, 50)}$=0.5, *P*=0.6. (**D**) **Intensity** ratings were different in $CS^-$ vs. $CS^+$ ($F_{(1, 50)}$=5.2, *P*=0.03), with a non-significant effects for Group ($F_{(2, 50)}$ = 0.1, *P*=0.9) nor for interaction ($F_{(2, 50)}$=0.9, *P*=0.4). (**E**) **Pleasantness** ratings, however, did not differ according to stimulus ($F_{1, 50)}$=0.5, *P*=0.5) nor according to group ($F_{(1, 50)}$=0.7, *P*=0.5; Interaction: $F_{(2, 50)}$= 2.5, *P*=0.1). **(F) Intake** was similar across conditioning for $CS^-$ and $CS^+$ flavours ($F_{(1, 50)}$=0.5, *P*=0.5) and despite findings of a significant group effect ($F_{(1, 50)}$=7.5, *P*=0.001), interaction between factors was not significant ($F_{(2, 50)}$=1.0, *P*=0.4; mixed-model 2-way ANOVA).

From pre- to post-conditioning days, ratings remained stable, and did not vary according to group for **(G) Hunger** (Time: $F_{(1, 47)}$=0.2, *P*=0.6; Group: $F_{(2, 50)}$=1.98, *P*=0.2; Interaction: $F_{(2, 47)}$=0.3, *P*=0.7; and **(H) Thirst** (Time: F $_{(1, 47)}$=0.05, *P*=0.8; Group: $F_{(2, 50)}$=2.5, *P*=0.1; Interaction: $F_{(2, 47)}$=0.8; *P*=0.5). **(I) Novelty** ratings changed from pre to post-conditioning ($F_{(1, 50)}$=12.9, *P*=0.001), with a significant effect for group ($F_{(2, 50)}$=3.4, *P*=0.04) and a non-significant interaction between factors ($F_{(2, 50)}$=0.7; *P*=0.5). Post-hoc tests showed significant decreases for the surgical group (*P*=0.05), while in the remaining groups results did not reach significance (Healthy, *P*=0.4; Obese, *P*=0.1). **(J) Intensity** ratings remained similar from pre to post-conditioning ($F_{(1, 50)}$= 0.7, *P*=0.4), with no effects for group ($F_{(2, 50)}$ = 0.66, *P*=0.52) nor interaction ($F_{(2, 50)}$=1.5; *P*=0.2; mixed-model 2-way ANOVA).

Bar graphs represent mean ± standard error of the mean (SEM).
gLMS/ gLHS - general labeled magnitude/hedonic scale.
VAS: Visual Analogue Scale.
*P ≤ 0.05; **P ≤ 0.01; ***P ≤ 0.001

**Supplementary Table 1. Demographic characteristics of healthy subjects.**

| Variable | Healthy 'FNC development' protocol | | | |
|---|---|---|---|---|
| | All (n=52) | Baseline low intake %pref. $CS^+$ (n=28)[A] | Baseline high intake %pref. $CS^+$ (n=24)[A] | *P*-value[C] |
| | Mean (SD), Min-Max or No. (%) | | | |
| Age, years | 28.5 (7.1), 18 – 48 | 28.46 (7.25), 18 – 47 | 28.58 (7.17), 20 – 48 | 0.95 |
| Gender (male) | 15 (28.8%) | 7 (25.0%) | 8 (33.3%) | 0.51 |
| BMI, $Kg/m^2$ | 23.1 (3.2), 18.0 – 34.0 | 22.92 (2.57), 19.08 – 31.16 | 23.22 (3.88), 17.99 – 34.01 | 0.74 |
| Education, years | 14.2 (2.6), 9 – 19 | 13.71 (2.66), 9 – 19 | 14.83 (2.46), 12 – 17 | 0.12 |
| Smokers | 12 (23.1%) | 7 (11.1%%) | 5 (20.8%) | 0.72 |

[A]Baseline low intake preference for $CS^+ < 50\%$ in the pre-test; [B]Baseline high intake preference for $CS^+ \geq 50\%$ in the pre-test.

[C]Independent samples t-tests were performed for continuous variables and $\chi 2$ tests for categorical variables for comparisons between baseline low and high intake preference for $CS^+$

**Supplementary Table 2. Gustatory and psychometric measures of feeding behaviour in healthy subjects.**

| Variable | Healthy ('FNC development' protocol) | | | |
|---|---|---|---|---|
| | All (n=52) | Baseline low intake %pref. $CS^+$ (n=28)[A] | Baseline high intake %pref. $CS^+$ (n=24)[B] | *P* value[C] |
| | Mean (SD), Min-Max or No. (%) | | | |
| Taste Tresholds, *dB* | 4.27 (8.75), -6 – 34.0 | 2.84 (7.78), -6 – 24 | 5.78 (9.65), -6 – 34 | 0.31 |
| Accuity | 13.14 (2.49), 7.00 – 16.0 | 12.8 (2.5), 7 – 16 | 13.5 (2.48), 7 -16 | 0.38 |
| Sour ratings, *mm* | | | | |
| Intensity | 57.11 (19.76), 20.00 – 98.0 | 56.26 (18.7), 20 – 97.5 | 58.13 (21.41), 27 – 98.0 | 0.76 |
| Pleasantness | -33.71 (32.55), -94.75 – 41.5 | -34.61 (31.62), -81 – 41.5 | -32.63 (34.43), -94.75 – 29.25 | 0.84 |
| Salt ratings, *mm* | | | | |
| Intensity | 28.38 (14.56), 5.75 – 72.75 | 26.24 (11.71), 11 – 51.25 | 30.95 (17.34), 5.75 – 72.75 | 0.29 |
| Pleasantness | -4.59 (18.70), -76.0 – 25.75 | -2.47 (16.13), -43 – 25.75 | -7.13 (21.54), -76 – 15.75 | 0.42 |
| Sweet ratings, *mm* | | | | |
| Intensity | 17.05 (8.0), 6.25 – 38.50 | 16.07 (6.90), 6.25 – 30.50 | 18.23 (9.19), 6.75 – 38.50 | 0.38 |
| Pleasantness | 8.97 (9.73), -16-75 – 34.50 | 7.48 (9.73), -16.75 – 27.75 | 10.75 (9.67), -5.25 – 34.50 | 0.27 |
| Bitter ratings, *mm* | | | | |
| Intensity | 40.22 (20.15) 6.25 – 79.50 | 42.58 (21.26), 6.25 – 78.75 | 37.38 (18.86), 16.25 – 79.50 | 0.4 |

**Supplementary Table 2. (*Continued*)**

| Variable | All (n=52) | Baseline low intake pref. $CS^+$ (n=28)[A] | Baseline high intake pref. $CS^+$ (n=24)[B] | *P* value[C] |
|---|---|---|---|---|
| | **Healthy ('FNC development' protocol)** | | | |
| | **Mean (SD), Min-Max or No. (%)** | | | |
| Pleasantness | -36.23 (24.02),-74.25 – -1.50 | -39.46 (25.71), -74.25 – -1.50 | -32.35 (21.82), -72.25 – -6.0 | 0.33 |
| PFS – Aggregate score | 2.28 (0.66), 1.13 – 3.80 | 2.23 (0.65), 1.13 – 3.67 | 2.35 (0.68), 1.47 – 3.80 | 0.51 |
| YFAS – Diagnosis | 0 (%) | 0 (%) | 0 (%) | - |
| YFAS – No. of symptoms | 1.54 (1.09), 0 – 5 | 1.46 (1.04), 0 – 4 | 1.64 (1.18), 0 – 5 | 0.59 |
| DEBQ – External Eat | 2.77 (0.53), 1.5 – 4.30 | 2.66 (0.49), 1.5 – 3.60 | 2.92 (0.56), 2.20 – 4.30 | 0.08 |
| DEBQ – Restrained Eat | 2.35 (0.74), 1.00 – 4.50 | 2.28 (0.64), 1.3 – 4 | 2.42 (0.87), 1.0 – 4.50 | 0.51 |
| DEBQ – Emotional Eat | 1.99 (0.76), 0.77 – 4.38 | 1.81 (0.64), 0.77 – 3.15 | 2.21 (0.84), 1.0 – 4.38 | 0.06 |
| FARS – Aggregate | 401.87 (43.08), 303.0 – 506.0 | 399.46 (48.37), 303.0 – 506.0 | 404.39 (37.72), 332.0 – 471.0 | 0.70 |

[A]Baseline low intake preference for $CS^+$ <50% in the pre-conditioning; [B]Baseline high intake preference for $CS^+ \geq 50\%$ in the pre-conditioning.
[C]Independent samples t-tests were performed for continuous variables and $\chi 2$ tests for categorical variables for comparisons between baseline low and high intake preference for $CS^+$.
DEBQ - Dutch Eating Behaviour Questionnaire; FARS - Food Action Rating Scale; PFS - Power of Food Scale; YFAS - Yale Food Addiction Scale.

**Supplementary Table 3. Associations between striatal dopamine DD2lR availability, the conditioning strength, and feeding behaviour.**

| Striatal DD2lR availability | Healthy (n=18) | | Obese (n=9) | | Surgical | | | | | |
|---|---|---|---|---|---|---|---|---|---|---|
| | | | | | All (n=20) | | Bypass (n=11) | | Sleeve (n=9) | |
| | **r** | ***P*-value** | **r** | ***P*-value** | **r** | ***P*-value** | **r** | ***P*-value** | **r** | ***P*-value** |
| BMI, kg/m$^2$ | 0.23 | 0.36 | -0.78 | 0.01 | -0.30 | 0.19 | -0.05 | 0.89 | -0.49 | 0.18 |
| Δ intake pref. CS$^+$ | 0.04 | 0.87 | 0.05 | 0.89 | -0.14 | 0.57 | -0.68 | 0.02 | 0.49 | 0.18 |
| Taste Tresholds | -0.29 | 0.24 | -0.25 | 0.52 | -0.05 | 0.86 | 0.07 | 0.85 | -0.18 | 0.70 |
| Acuity | -0.20 | 0.42 | -0.40 | 0.28 | -0.14 | 0.55 | 0.28 | 0.41 | -0.50 | 0.17 |
| Sour ratings (mm) | | | | | | | | | | |
| Intensity | -0.20 | 0.42 | -0.40 | 0.28 | -0.14 | 0.55 | 0.28 | 0.41 | -0.50 | 0.17 |
| Pleasantness | 0.18 | 0.48 | 0.26 | 0.49 | 0.11 | 0.66 | 0.21 | 0.53 | 0.02 | 0.96 |
| Salt ratings (mm) | | | | | | | | | | |
| Intensity | 0.05 | 0.83 | -0.61 | 0.08 | -0.38 | 0.10 | -0.29 | 0.39 | -0.44 | 0.24 |
| Pleasantness | -0.08 | 0.76 | 0.65 | 0.06 | 0.05 | 0.85 | 0.38 | 0.24 | -0.32 | 0.41 |
| Sweet ratings (mm) | | | | | | | | | | |
| Intensity | 0.06 | 0.82 | -0.44 | 0.23 | -0.36 | 0.11 | -0.46 | 0.15 | -0.27 | 0.49 |

**Supplementary Table 3. (C*ontinued*)**

| Striatal DD2lR availability | Healthy (n=18) | | Obese (n=9) | | Surgical | | | | | |
|---|---|---|---|---|---|---|---|---|---|---|
| | | | | | All (n=20) | | Bypass (n=11) | | Sleeve (n=9) | |
| | r | *P*-value | r | *P*-value | r | *P*-value | r | *P*-value | r | *P*-value |
| Pleasantness | -0.06 | 0.80 | -0.03 | 0.94 | 0.19 | 0.42 | -0.04 | 0.91 | 0.55 | 0.12 |
| Bitter ratings (mm) | | | | | | | | | | |
| Intensity | -0.06 | 0.80 | -0.28 | 0.47 | -0.34 | 0.14 | -0.55 | 0.08 | -0.06 | 0.88 |
| Pleasantness | 0.11 | 0.67 | 0.19 | 0.63 | 0.30 | 0.21 | 0.46 | 0.16 | 0.09 | 0.81 |
| PFS – Aggregate score | -0.02 | 0.93 | -0.63 | 0.07 | 0.13 | 0.60 | 0.17 | 0.61 | 0.13 | 0.76 |
| PFS – Food Available | 0.03 | 0.90 | -0.58 | 0.10 | 0.15 | 0.55 | 0.41 | 0.20 | -0.14 | 0.74 |
| PFS – Food Present | 0.14 | 0.57 | -0.55 | 0.12 | -0.10 | 0.69 | -0.33 | 0.32 | 0.15 | 0.73 |
| PFS – Food tasted | -0.25 | 0.32 | -0.61 | 0.08 | 0.22 | 0.36 | 0.12 | 0.74 | 0.39 | 0.34 |
| YFAS – No. of symptoms | -0.22 | 0.39 | -0.61 | 0.08 | -0.17 | 0.49 | -0.11 | 0.76 | -0.16 | 0.68 |
| DEBQ – External Eat | -0.12 | 0.64 | -0.46 | 0.21 | 0.16 | 0.51 | 0.36 | 0.28 | -0.21 | 0.61 |
| DEBQ – Restrained Eat | 0.16 | 0.52 | 0.28 | 0.47 | 0.19 | 0.44 | 0.77 | 0.01 | -0.26 | 0.54 |
| DEBQ – Emotional Eat | 0.16 | 0.53 | -0.66 | 0.05 | -0.22 | 0.39 | 0.16 | 0.65 | -0.57 | 0.18 |
| FARS – Aggregate | -0.29 | 0.25 | -0.12 | 0.77 | 0.03 | 0.91 | 0.26 | 0.47 | -0.23 | 0.58 |

BMI - Body Mass Index; DEBQ - Dutch Eating Behaviour Questionnaire; PFS - Power of Food Scale; FARS - Food Action Rating Scale; Striatal DD2lR availability – Striatal dopamine D2-like receptor availability; YFAS - Yale Food Addiction Scale. Δ intake pref. $CS^+$ (i.e., 'the conditioning strength'): the difference between intake preference for $CS^+$ (%) in the post minus the pre-test.
